# Pan-cancer analyses reveal genomics and clinical characteristics of the down regulated genes for recurrent myocardial infarction in cancer

**DOI:** 10.1101/2022.02.21.22271285

**Authors:** Zongyu Wang

**Affiliations:** Department of Traditional Chinese Medicine, Guangxi Medical University Cancer Hospital, Nanning 530021, China

**Keywords:** Neoplasms, Myocardial Infarction, Genomics, GEO, Prognosis

## Abstract

**Backgroun:** Early identification of the occurrence and progression of coronary ischemic events is particularly important for the diagnosis and treatment of coronary heart disease complicated with tumor. Therefore, it is of great significance to analyze biomarkers and regulatory factors of recurrent myocardial infarction after first myocardial infarction.In order to specify the regulatory factor of coronary heart disease or myocardial infarction on tumor occurrence and metastasis and clinical survival effect, we select from GEO database for the first time of recurrence after myocardial infarction, myocardial infarction, microarray data set, the comprehensive evaluation of the data set is the key cut genes and cancer, tumor prognosis, the tumor extends into the immune level of mutation, etc.The purpose of this study is to discover the role of myocardial infarction regulatory factors in cancer.

**Methods:** GSE48060 chip data set was downloaded from GEO database to obtain gene expression profiles of blood samples from normal cardiac function control and first AMI patients within 48 hours after the first myocardial infarction.GEO2R online tool and Excel software were used to screen the differential genes between the reinfarction samples and the normal samples in the GSE48060 chip data set.DAVID database was used for enrichment analysis of GO and KEGG of DEGs.The PPI network was constructed and visualized by Using The STRING database and Cytoscape software, and the key genes were screened by the CytoHubba plug-in of Cytoscape software.TIMER database and GEPIA2 website were used to analyze and verify the expression levels of key genes between tumor tissues and normal tissues in generalized carcinoma.GEPIA2 website was used to analyze the correlation between key genes and tumor prognosis.The correlation between key gene expression and tumor purity and each immune cell was analyzed by TIMER database.CBioPortal website was used to analyze the mutation and correlation of key genes in tumors.

**Results:** Through analysis, the difference genes of 17 GSE48060 chip data sets of reinfarction samples and normal samples were obtained, and the above genes were all down-regulated genes.Five key genes were obtained through constructing PPI network, which were GART, SOD2, KLRF1, ERAP2 and CXCL5.According to the TIMER result, The expression level of GART in BLCA, BRCA, CHOL, COAD, ESCA, HNSC, KICH, KIRC, LIHC, LUAD, LUSC, PRAD, READ, STAD and THCA was significantly different from that in normal tissues (P < 0.001).The expression levels of SOD2 in BRCA, ESCA, KICH, KIRC, KIRP, LUSC, SKCM and STAD were significantly different from normal tissues (P < 0.001).The expression levels of KLRF1 in BRCA, COAD, HNSC, LIHC, LUAD, LUSC, PRAD, READ, STAD and UCEC were significantly different from those in normal tissues (P < 0.001).The expression levels of ERAP2 in BRCA, KIRC and LUSC were significantly different from those in normal tissues (P < 0.001).The expression levels of CXCL5 in BRCA, COAD, ESCA, HNSC, KICH, KIRC, KIRP, LUAD, LUSC, PRAD, STAD, THCA and UCEC were significantly different from those in normal tissues (P < 0.001).The results of GEPIA2 showed that the expression levels of GART in CESC, CHOL, ESCA, KICH, LUSC, SARC and STAD showed significant differences between tumor tissues and normal tissues (P < 0.05).There were significant differences in the expression levels of SOD2 in BLCA, BRCA, ESCA, PAAD and PCPG between tumor tissues and normal tissues (P < 0.05), and there were significant differences in the expression levels of KLRF1 in LUAD and LUSC between tumor tissues and normal tissues (P < 0.05).There were significant differences in ERAP2 expression between tumor tissues and normal tissues in KICH and LUSC (P < 0.05), and there were significant differences in CXCL5 expression between tumor tissues and normal tissues in CHOL, COAD, ESCA, LUAD, LUSC, READ and STAD (P < 0.05).The results of tumor survival analysis indicated that the correlation between the overall tumor survival (OS) of GART and CESC, KIRC, LUAD, READ and SARC was statistically significant (P < 0.05).The correlation between SOD2 and TUMOR OS of SKCM was statistically significant (P < 0.05).The correlation of TUMOR OS between KLRF1 and HNSC was statistically significant (P < 0.05).The correlation between CXCL5 and TUMOR OS of KICH and KIRC was statistically significant (P < 0.05).There was statistically significant correlation between GART and LUAD’s tumor stage (P < 0.05), SOD2 and SKCM’s tumor stage (P < 0.05), CXCL5 and KICH and KIRC’s tumor stage (P < 0.05).The GART gene expression level is significantly correlated with various immune cells in CESC, KIRC, LUAD, READ and SARC.The expression level of SOD2 gene was significantly correlated with various immune cells in SKCM.The expression level of KLRF1 gene was significantly correlated with various immune cells in HNSC.The expression level of CXCL5 gene was significantly correlated with various immune cells in KICH and KIRC.The proportion of mutations in COAD, KICH, LIHC, READ, SKCM and THCA was the highest, the proportion of amplification in BRCA, ESCA, HNSC, KIRC, LUSC and STAD was the highest, and the proportion of severe deletion in LUAD and PRAD was the highest.

**Conclusion:** The key down-regulated genes of recurrent myocardial infarction after initial myocardial infarction are related to tumor genesis, metastasis, clinical survival and immune invasion.Key down-regulated genes in recurrent myocardial infarction may serve as important biomarkers for the diagnosis and treatment of cancer.

## background

The rise of oncology cardiology in recent years has pointed the prognostic factors of oncology patients more toward the cardiovascular direction of [1].The respective risk factors and prognosis of malignant tumors and coronary heart disease have been extensively deeply studied. The common biological mechanism is inflammation, which can promote the malignant degeneration and progression of cells in the tumor microenvironment.But when the two diseases together, especially in the tumor progression or cardiovascular emergency, determine patients near / long prognosis risk factors are more complex, the pathophysiological state of the two not only affect each other, and treatment decisions often restrict each other, even lead to patients in a disease direction of treatment interruption, eventually lead to poor prognosis [2].At present, there are no relevant guidelines that can systematically standardize and guide the clinical monitoring and treatment of patients with CAD and tumors, nor is there a large amount of reliable evidence to reflect the diagnosis and treatment and prognosis of such patients.Among them, the early identification of the occurrence and progression of coronary ischemic events is particularly important for the diagnosis and treatment of coronary heart disease combined with tumors, so it is of great significance to analyze the biomarkers and regulators of recurrent MI after the first myocardial infarction.In order to clarify whether the regulators of coronary heart disease or myocardial infarction will have an impact on tumor development, metastasis and clinical survival, we selected from the GEO database recurrence after myocardial infarction chip dataset, comprehensively evaluated the key downregulated genes and pan-cancer, tumor prognosis, tumor immune infiltration level, mutations, etc.It aims to discover the role of myocardial infarction regulators in cancer through this study.

## 1. Materials and Methods

### 1.1 Data acquisition

The National Center for Biotechnology (Gene Expression Omnibus, GEO) [3] is a public database of storage chips, second-generation sequencing, and other high-throughput sequencing data.The microarray dataset numbered GSE48060[4] was downloaded from the GEO database to obtain gene expression profiles of blood samples from normal cardiac function controls and first AMI patients within 48 h after the first MI.The platform for GSE48060 was GPL570, including 21 normal control samples and 31 first MI samples, five of which had MI recurrence during the follow-up and 18 months after MI and 26.

### 1.2 Differential genetic screen

Applying the GEO2R (http://www.ncbi.nlm.nih.The gov / geo / geo2r) online tool and normal samples in the GSE48060 microarray data aset with screening criteria of P value <0.01, | logFC |> 1.

### 1.3 Enriched functional annotations of the differential genes

Import the DEGs into the DAVID 2021 version (https://david-d.ncifcrf.The gov) database was analyzed by Gene Ontology (Gene Ontology, GO) and the Kyoto Encyclopedia of Genes (Kyoto Encyclopedia of Genes and Genomes, KEGG), with P <0.05 considered statistically significant.

### 1.4 protein proteininteraction (PPI) network construction

By the STRING (https://www.string-db.The org /) database constructed a PPI network for DEGs, analyzed protein interactions, “Settings” set scores> 0.15 and saved in TSV format, subsequently import into Cytoscape 3.8.2 software optimization and visualization of the PPI network.

### 1.5 Screening of the key genes

The CytoHubba plugin was used to screen key genes, CytoHubba plug-in according to the properties of nods in the network, high degree proteins are more inclined to key protein [5], MCC is a new proposed method, in PPI network, MCC analysis, integrated degree and MCC algorithm selected TOP5 genes as key genes.

### 1.6 Expression analysis of key genes in pan-cancer

Apply the TIMER database (https://cistrome.shinyapps.For io / TIMER /) analysis of the expression levels of key genes in pan tumor versus normal tissues, P <0.001 was considered statistically significant.Reapply the GEPIA2 website (http://gepia2.cancer-pku.The cn / # index) validate the expression levels of key genes in pan tumor versus normal tissues, P <0.05 was considered statistically significant, and Matched normal data selected match TCGA normal data.

### 1.7 Correlation analysis of key genes and tumor prognosis

Key genes were imported into GEPIA2, selected tumor diseases with significant differences in gene expression levels between tumor and normal tissues, performed overall survival survival analysis (OS), and drew Kaplan-Meier survival curves, with P <0.05 considered statistically significant.Reapplying GEPIA2 to analyze the correlation of key genes and tumor stage, P <0.05 was considered statistically significant.

### 1.8 Correlation analysis of key gene expression and the level of immune infiltration in tumors

Key genes were imported into the TIMER database, and tumor diseases with significant correlation with the tumor prognosis of each key gene were selected for the correlation analysis between key gene expression and tumor purity and each immune cell.

### 1.9 Mutation and correlation analysis of key genes in tumors

Apply the cBioPortal website (https://www.cbioportal.To org /), analyze the mutations and correlations of key genes in tumors, and observe the proportion of mutations, amplification, and severe deletions in different tumors.

## 2. Results

### 2.1 Identification and functional annotation of DEGs

The GSE48060 microarray data set yielded 17 DEGs, all of which were downregulated genes (Figure 1).Functional analysis of the DEGs was performed using the DAVID.As shown in Table 1, the results indicate that DEGs are mainly enriched in response to axonal damage, synaptic vesicle endocytosis, protein homo-tetramer, adaptive immune response, blood pressure regulation.

**Figure 1.**
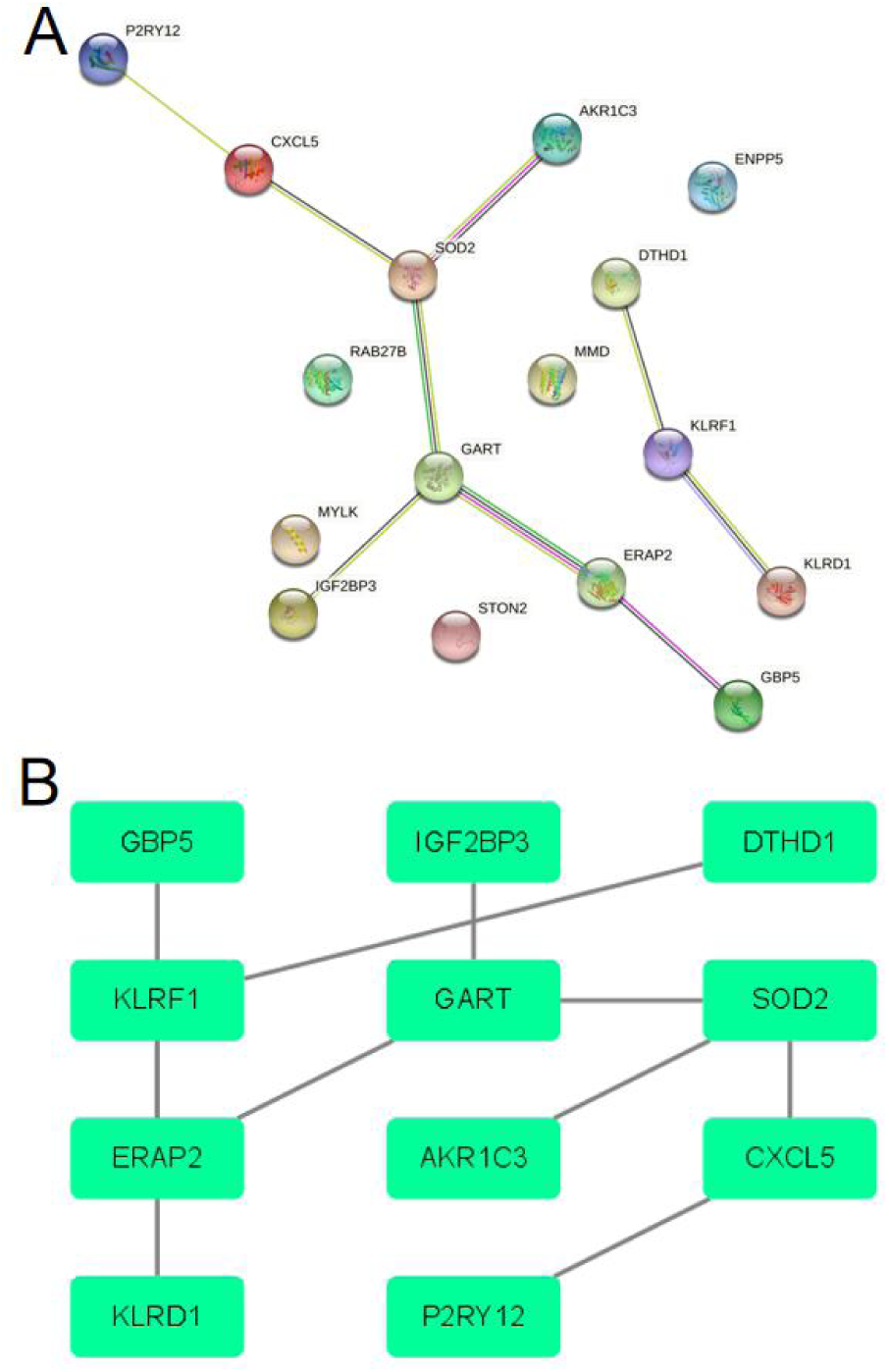
PPI network diagram of DEGs. A.The PPI network built by the STRING database; B.Cytoscape software optimized and visualized the PPI networks

**Table 1.** Enrichment analysis of the GO and KEGG pathway for D E G s.

| clauses and subclauses | description | count | percentage (%) | P value |
| --- | --- | --- | --- | --- |
| GO:0048678 | response to axon injury | 2 | 11.76470588 | 0.028563638 |
| GO:0048488 | synaptic vesicle endocytosis | 2 | 11.76470588 | 0.034183792 |
| GO:0051289 | protein homotetramerization | 2 | 11.76470588 | 0.046124496 |
| GO:0002250 | adaptive immune<br>response | 3 | 17.64705882 | 0.052234624 |
| GO:0008217 | regulation of blood<br>pressure | 2 | 11.76470588 | 0.058708618 |

### 2.2 Construction of P P I networks and screening of key genes

DEGs were imported into the STRING database to build the PPI network, and then optimized and visualized by Cytoscape software, as shown in Figure 1. Using the CytoHubba plugin in Cytoscape software, the genes of TOP5 were selected as key genes, namely GART, SOD2, KLRF1, ERAP2, and CXCL5, whose full gene name and function are shown in Table 2.

**Table 2.**
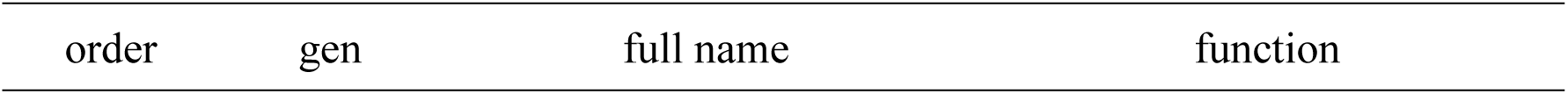
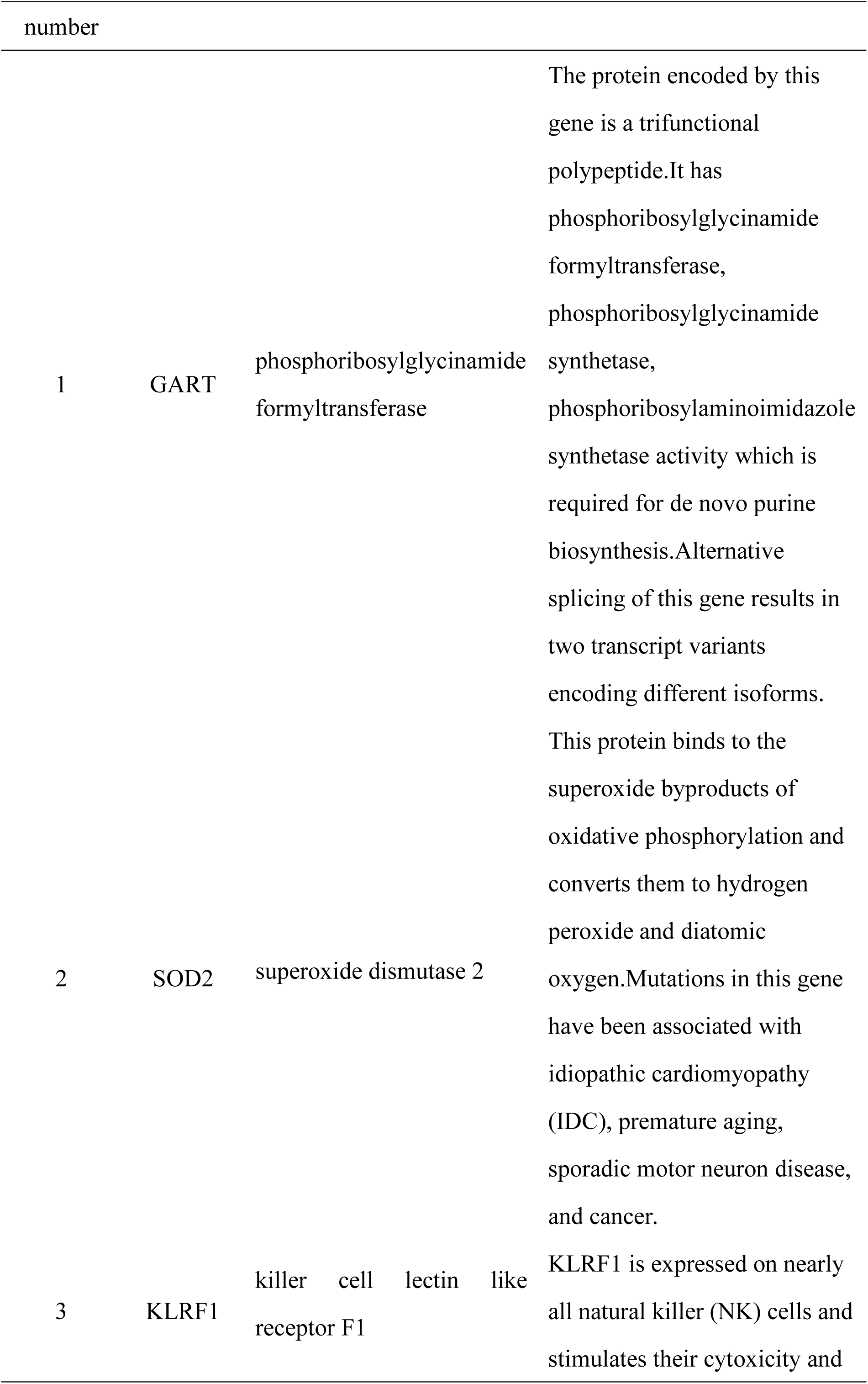
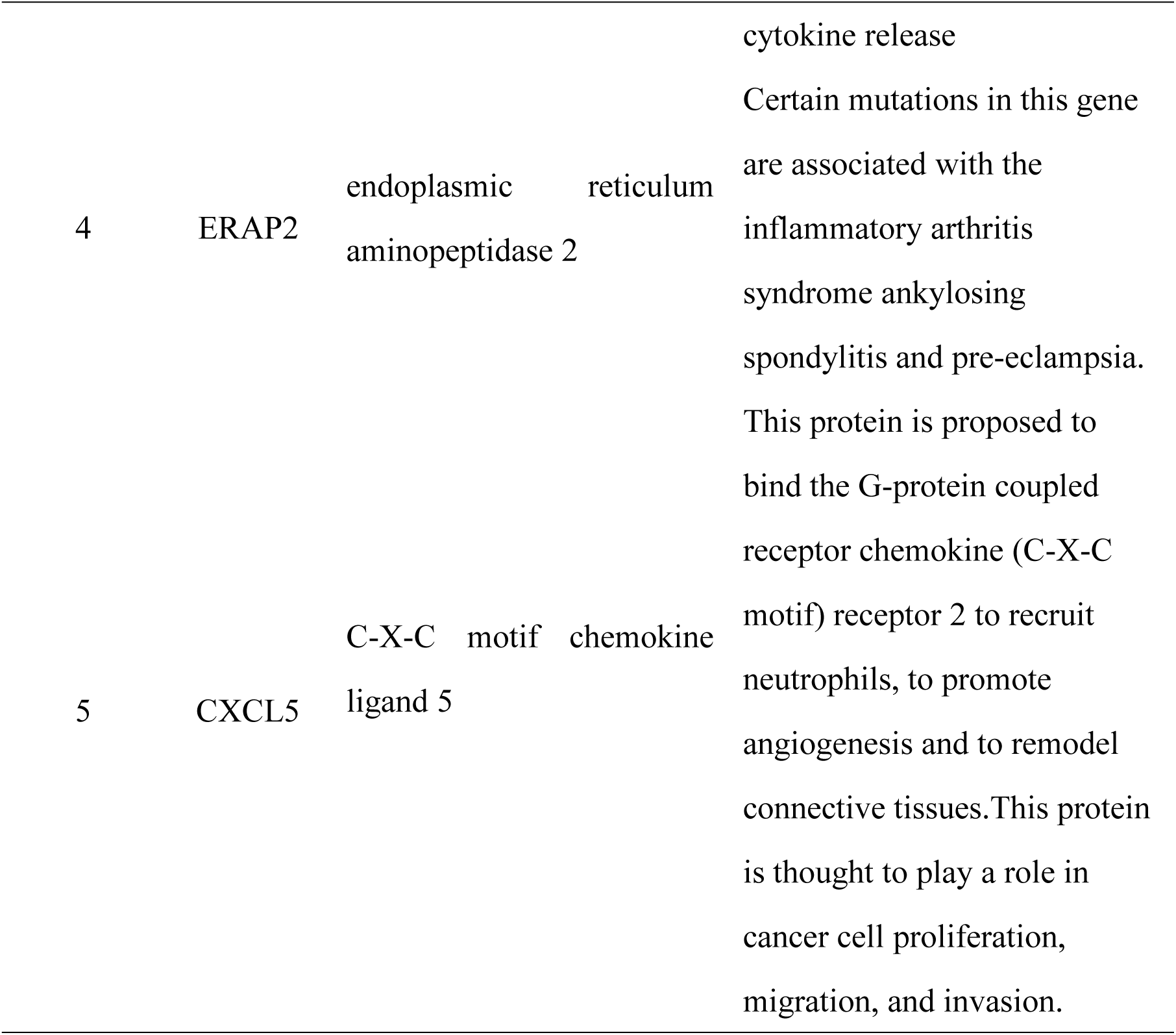
Names and functional roles of the key genes.

### 2.3 Expression analysis of key genes in pan-cancer

Five key genes were introduced into TIMER, and the expression levels of key genes in tumor tissues and normal tissues of generalized cancer were analyzed, as shown in Figure 2.The results showed that the expression level of GART in BLCA, BRCA, CHOL, COAD, ESCA, HNSC, KICH, KIRC, LIHC, LUAD, LUSC, PRAD, READ, STAD and THCA was significantly different from that in normal tissues (P < 0.001).The expression levels of SOD2 in BRCA, ESCA, KICH, KIRC, KIRP, LUSC, SKCM and STAD were significantly different from normal tissues (P < 0.001).The expression levels of KLRF1 in BRCA, COAD, HNSC, LIHC, LUAD, LUSC, PRAD, READ, STAD and UCEC were significantly different from those in normal tissues (P < 0.001).The expression levels of ERAP2 in BRCA, KIRC and LUSC were significantly different from those in normal tissues (P < 0.001).The expression levels of CXCL5 in BRCA, COAD, ESCA, HNSC, KICH, KIRC, KIRP, LUAD, LUSC, PRAD, STAD, THCA and UCEC were significantly different from those in normal tissues (P < 0.001).Then 5 key genes were introduced into GEPIA2 to verify the expression levels of key genes in the tumor tissues and normal tissues of generalized carcinoma. The results were shown in Figure 3.The expression levels of GART in CESC, CHOL, ESCA, KICH, LUSC, SARC and STAD showed significant differences between tumor tissues and normal tissues (P < 0.05).There were significant differences in the expression levels of SOD2 in BLCA, BRCA, ESCA, PAAD and PCPG between tumor tissues and normal tissues (P < 0.05), and there were significant differences in the expression levels of KLRF1 in LUAD and LUSC between tumor tissues and normal tissues (P < 0.05).There were significant differences in ERAP2 expression between tumor tissues and normal tissues in KICH and LUSC (P < 0.05), and there were significant differences in CXCL5 expression between tumor tissues and normal tissues in CHOL, COAD, ESCA, LUAD, LUSC, READ and STAD (P < 0.05).Five key genes were imported into the GEPIA2, Validate the expression levels of key genes in the tumor versus normal tissues of pan-cancer, The results are shown in Figure 3, The expression levels of GART in tumor tissues and normal tissues in CESC, CHOL, ESCA, KICH, LUSC, SARC, and STAD varied significantly (P <0.05), The expression levels of SOD2 in tumor tissues and normal tissues in BLCA, BRCA, ESCA, PAAD and PCPG (P <0.05), The expression levels of KLRF1 in tumor tissues and normal tissues in LUAD, LUSC were significantly different (P <0.05), The expression levels of ERAP2 in KICH and LUSC were significantly different (P <0.05), The expression levels of CXCL5 in CHOL, COAD, ESCA, LUAD, LUSC, READ, and STAD varied significantly (P <0.05).

**Figure 2.**
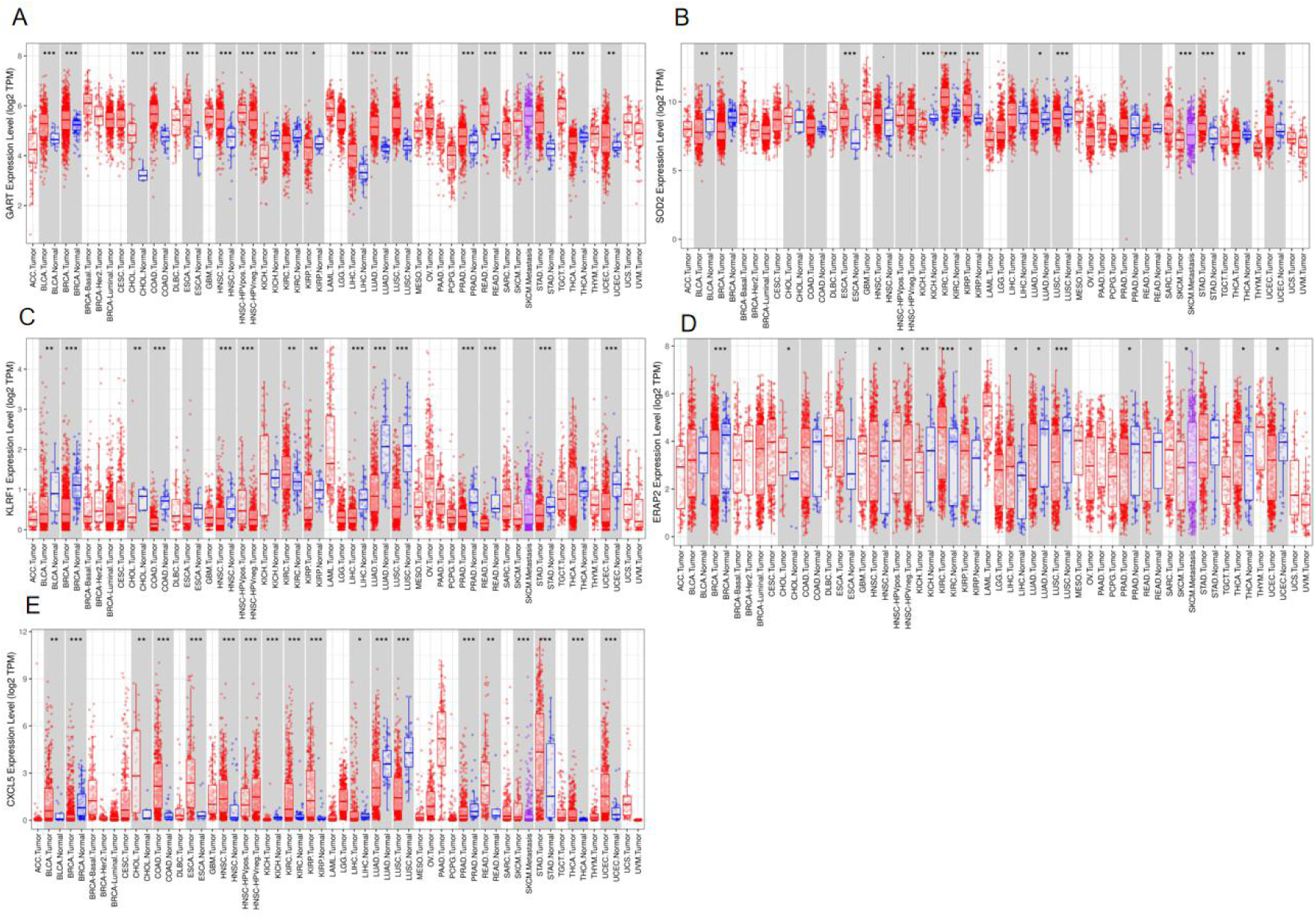
Expression levels of key genes in the tumor and normal tissues of pan-cancer (TIMER) A.Expression levels of GART in the tumor and normal tissues of pan-cancer; B.Expression levels of SOD2 in tumor and normal tissues; C.Expression levels of KLRF1 in tumor and normal tissues; D.Expression levels of ERAP2 in the tumor and normal tissues of pan-cancer; E.Expression levels of CXCL5 in the tumor and normal tissues of pan-cancer

**Figure 3.**
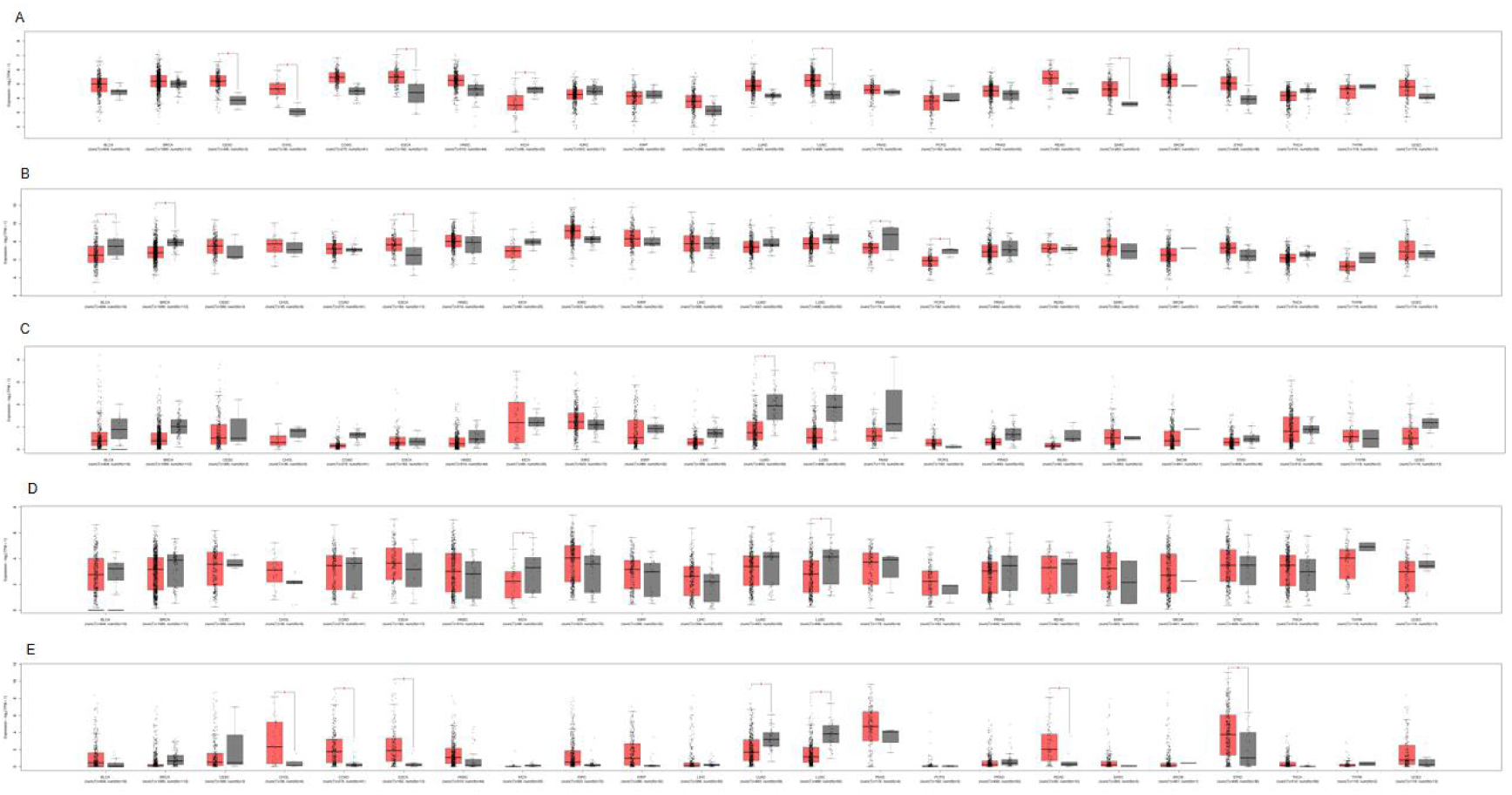
Expression levels of key genes in the tumor and normal tissues of pan-cancer (GEPIA2) A.Expression levels of GART in the tumor and normal tissues of pan-cancer; B.Expression levels of SOD2 in tumor and normal tissues; C.Expression levels of KLRF1 in tumor and normal tissues; D.Expression levels of ERAP2 in the tumor and normal tissues of pan-cancer; E.Expression levels of CXCL5 in the tumor and normal tissues of pan-cancer

### 2.4 Correlation analysis of key genes and tumor prognosis

Five key genes were introduced into GEPIA2, and tumor diseases with significant difference in gene expression level between tumor tissues and normal tissues in 2.3 were selected for survival analysis.Results As shown in Figure 4, the correlation between the overall tumor survival (OS) of GART and CESC, KIRC, LUAD, READ, and SARC was statistically significant (P < 0.05).The OS of those with high expression of GART in kirc and read was longer than that of those with low expression, and that of those with low expression of GART in CESC, LUAD and SARC was longer than that of those with high expression.The correlation between SOD2 and TUMOR OS of SKCM was statistically significant (P < 0.05), and the OS of those with high expression of SOD2 in SKCM was longer than that of those with low expression.The correlation of TUMOR OS between KLRF1 and HNSC was statistically significant (P < 0.05), and the patients with high and low KLRF1 expression in HNSC had longer OS.The correlation between CXCL5 and TUMOR OS of KICH and KIRC was statistically significant (P < 0.05),and the OS of those with low expression of CXCL5 in KICH and KIRC was longer than that of those with high expression.

**Figure 4.**
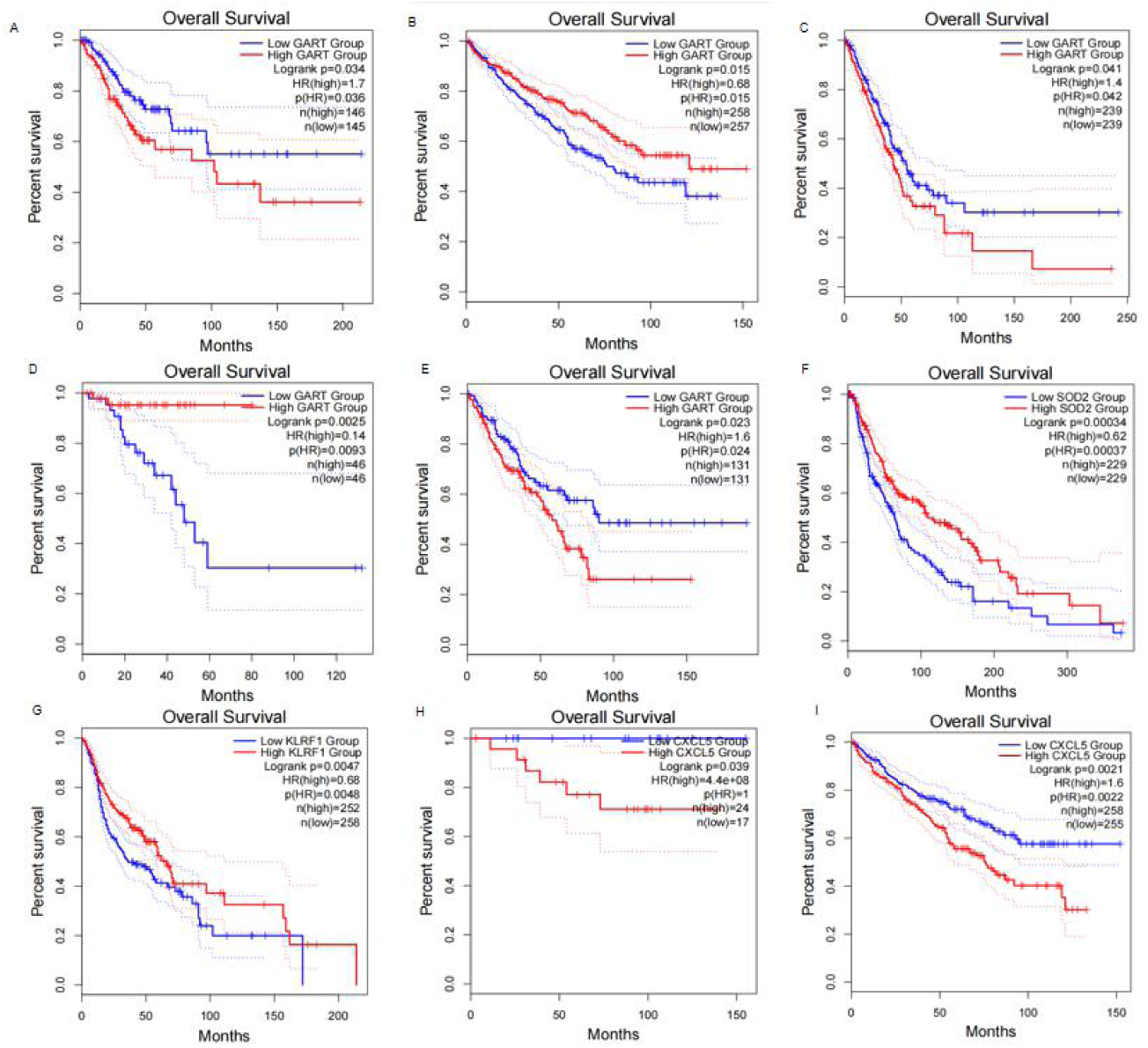
Association analysis of key genes and tumor OS. A.Correlation between the GART and the OS of the CESC; B.Correlation between GART and the OS of KIRC; C.Correlation between GART and the OS of LUAD; D.Correlation between GART and the OS of READ; E.Correlation between the GART and the OS of the SARC; F.Correlation of SOD2 and the OS of SKCM; G.Correlation of KLRF1 and the OS of HNSC; H.Correlation of CXCL5 and the OS of KICH; I.Correlation of CXCL5 and the OS of KIRC

The correlation between key genes and tumor stage was shown in Figure 5. The correlation between GART and LUAD’s tumor stage was statistically significant (P < 0.05), and that between SOD2 and SKCM’s tumor stage was statistically significant (P < 0.05).The correlation between CXCL5 and tumor stages of KICH and KIRC was statistically significant (P < 0.05).

**Figure 5.**
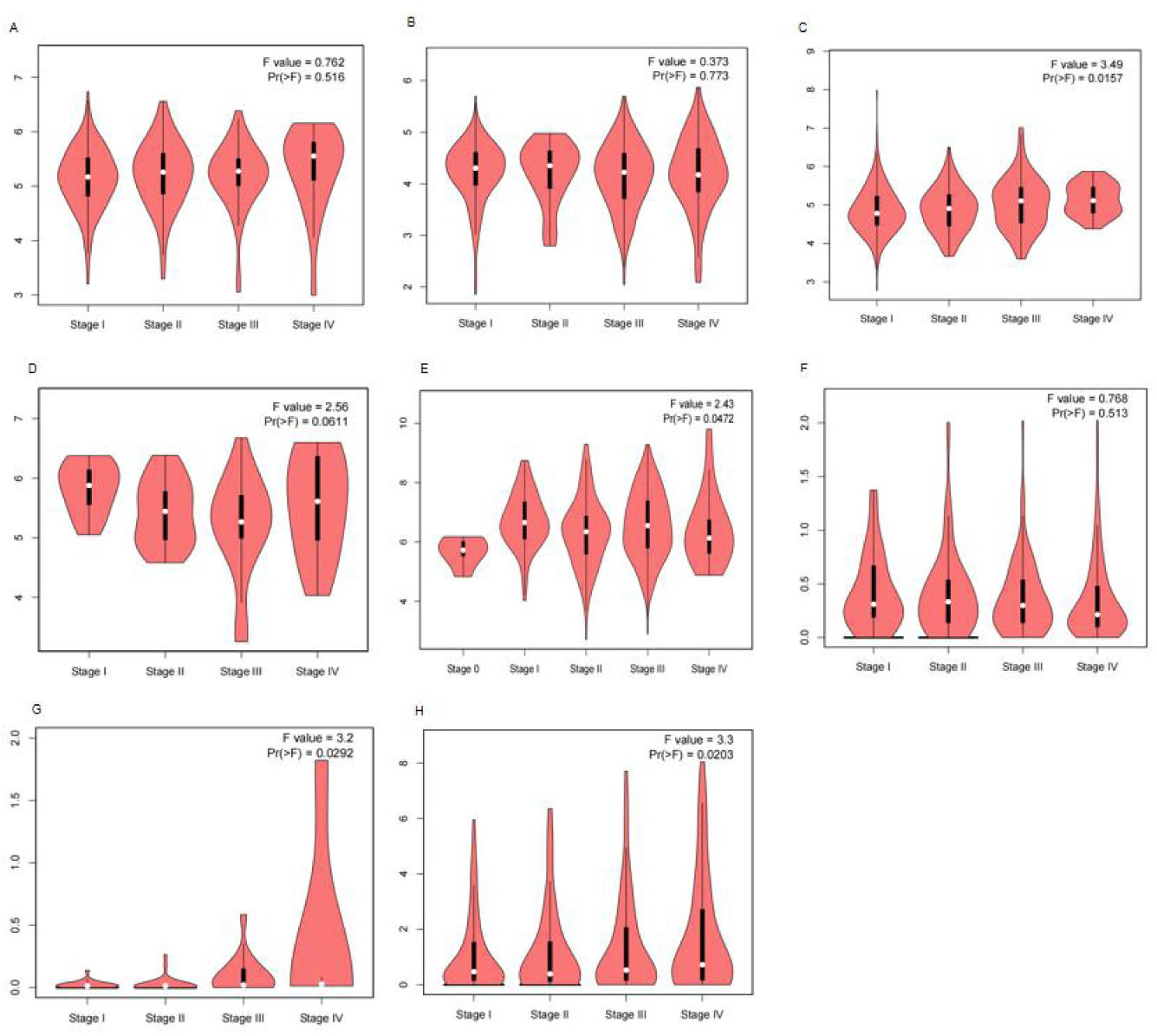
Correlation analysis of key genes and tumor stage. A.Correlation between GART and tumor stage of CESC; B.Correlation between GART and the tumor stage of KIRC; C.Correlation between GART and tumor stage of LUAD; D.Correlation between GART and tumor stage of READ; E.Correlation of SOD2 and the tumor stage of SKCM; F.Correlation of KLRF1 and tumor stage of HNSC; G.Correlation between CXCL5 and the tumor stage of KICH; H.Correlation of CXCL5 and tumor stage of KIRC

### 2.5 Correlation analysis of key gene expression and the level of immune infiltration in tumors

Independent tumor-infiltrating lymphocytes play a key role in predicting overall survival and sentinel lymph node status, [6].In order to study the relationship between key genes and multiple immune-infiltrating cells, we selected tumor diseases with significant correlation between tumor prognosis and key genes in the TIMER database, focusing on the correlation analysis between key gene expression and various immune cells (T cells (general), B cells, CD4 + T cells, CD8 + T cells, macrophages, neutrophils, dendritic cells).As shown in Figure 6, the results showed that GART gene expression level was significantly associated with various immune cells in CESC, KIRC, LUAD, READ, and SARC; SOD2 gene expression level was significantly associated with various immune cells in SKCM; KLRF1 gene expression level with various immune cells in HNSC; and CXCL5 gene expression level was significantly associated with various immune cells in KICH and KIRC.These findings suggest that key genes may modulate various immune cell polarization in the corresponding tumors.

**Figure 6.**
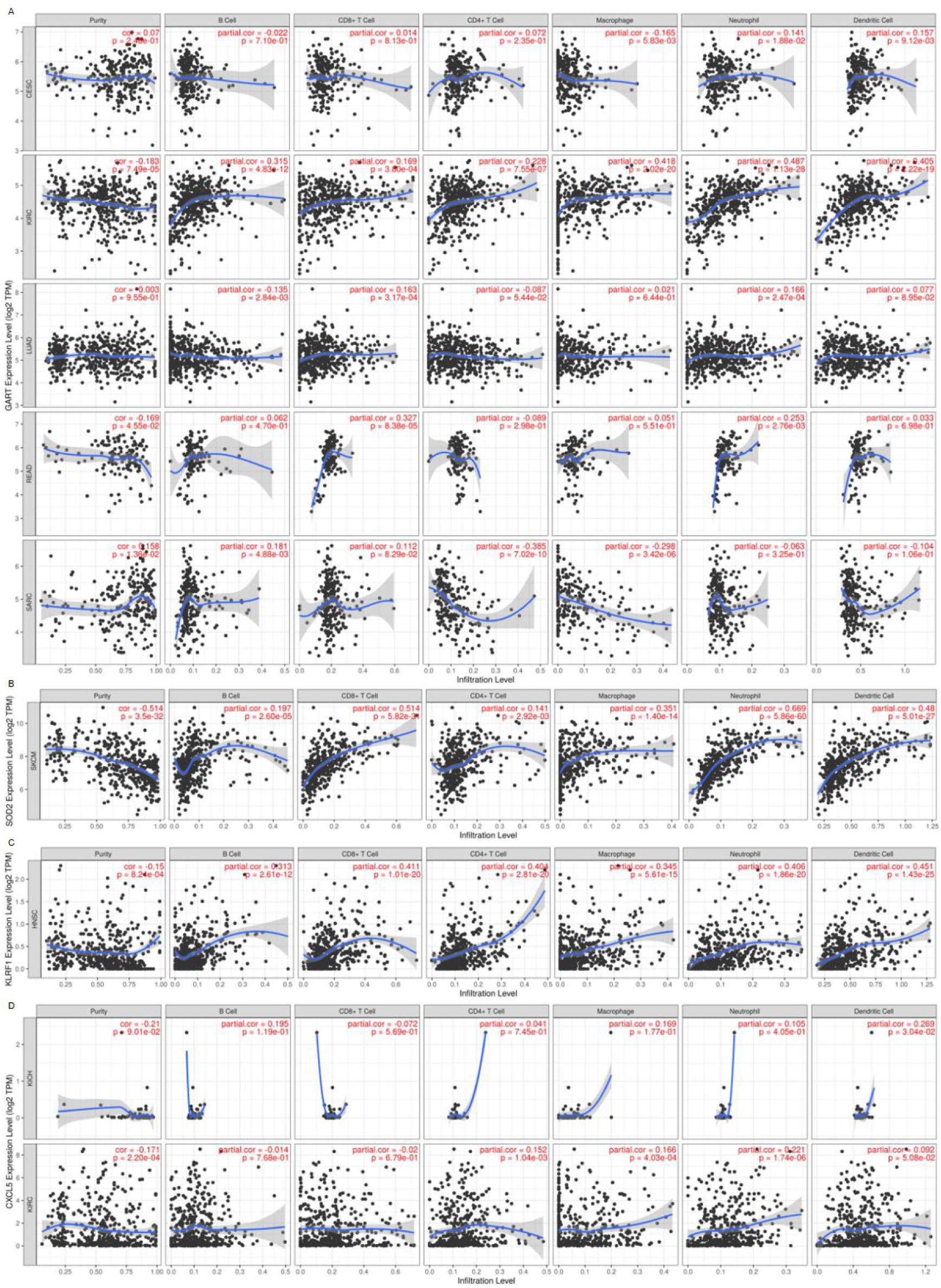
Correlation of key gene expression and the level of immune infiltration in tumors. A.Correlation between GART and the levels of immune infiltration in CESC; B.Correlation between GART and the levels of immune infiltration in KIRC; C.Correlation between GART and the levels of immune infiltration in LUAD; D.Correlation between GART and the levels of immune infiltration in READ; E.Correlation between GART and the levels of immune infiltration in SARC; F.Correlation between SOD2 and the levels of immune infiltration in SKCM; G.Correlation between KLRF1 and the level of immune infiltration in HNSC; H.Correlation between CXCL5 and the level of immune infiltration in KICH; I.Correlation between CXCL5 and the level of immune infiltration in KIRC

### 2.6 Mutation and correlation analysis of key genes in tumors

According to the expression levels of five key genes in Pan cancer tumor tissues and normal tissues, BRCA, coad, ESCA, HNSC, KICH, KIRH, LIHC, LUAD, LUSC, PRAD, READ, SKCM, STAD and THCA were introduced into cbioportal website to analyze the mutation and correlation of five key genes in the above tumors. The results are shown in Fig. 7, in which the mutation proportion is the largest in COAD, KICH, LIHC, READ, SKCM and THCA, the amplification proportion is the largest in BRCA, ESCA, HNSC, KIRC, LUSC and STAD, and the serious deletion proportion is the largest in LUAD and PRAD.

**Figure 7.**
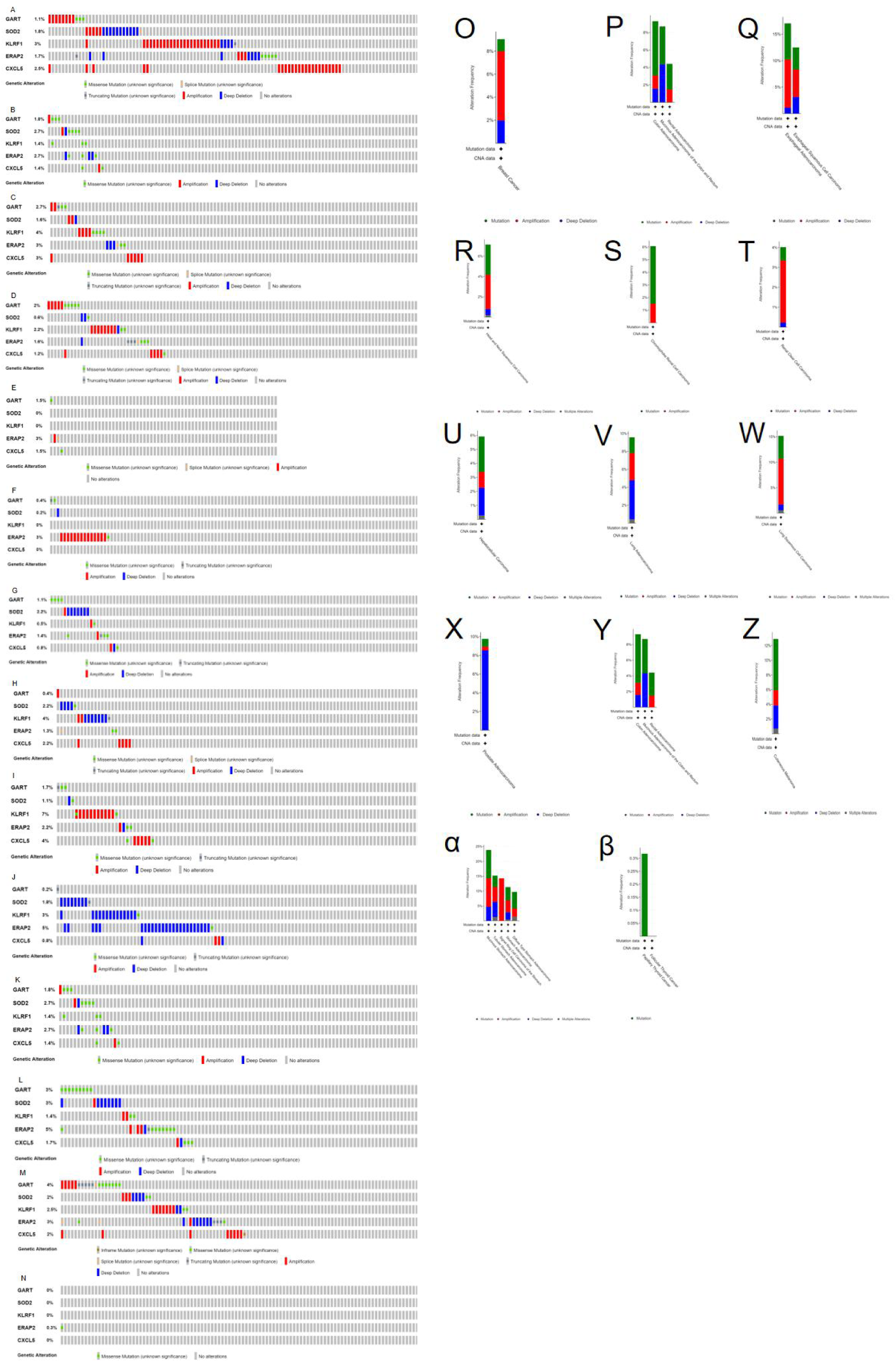
Mutation and correlation analysis of key genes in tumors. A.O.Mutations in the key gene, BRCA; B.P.Mutations of key genes in COAD; C.Q.Mutations of key genes in ESCA; D.R.Mutations of key genes in HNSC; E.S.Mutations of key genes in KICH; F.T.Mutations of key genes in KIRC; G.U.Mutations of key genes in LIHC; H.V.Mutations of key genes in LUAD; I.W.Mutations of key genes in LUSC; J.X.Mutations of key genes in PRAD; K.Y.Mutations of key genes in READ; L.Z.Mutations of key genes in SKCM; M.α.Mutations of key genes in STAD; N.β.Mutations of key genes in THCA

## 3. Discussion

At present, domestic and foreign studies have proved that there is a certain link between coronary heart disease and tumor incidence and survival.There are two foreign studies show that in acute myocardial infarction and percutaneous coronary intervention therapy (Percutaneous Coronary Intervention, PCI) of coronary heart disease patients, the history of malignant tumor is 7.8% and 5.6%, respectively, compared with the same patients, they are older, more complicated, and less accept PCI or standard secondary preventive drug treatment [7,8].Landes et al showed that the common occurrence in tumor patients treated with PCI, besides colorectal and breast cancer, including male urinary tumors such as prostate and bladder cancer, was identical to [7,9], and another study by velders et al.In our study, the findings reveal not only the different underlying mechanisms of recurrent MI regulators in the cancer setting, but also that regulators of recurrent MI are associated with cancer pathways.In this study, we identified five key downregulated genes for relapse after the first MI, namely GART, SOD2, KLRF1, ERAP2, and CXCL5.

Our results show significant differences in the expression levels of tumor tissues and normal tissues of the five key downregulated genes in BLCA, BRCA, CHOL, COAD, ESCA, HNSC, KICH, KIRC, LIHC, LUAD, LUSC, PRAD, READ, STAD, THCA, and UCEC.The KLRF1 gene belongs to the family of killer cell lectin-like receptors (KLR), with KLRF1 mRNA detectable in NK cells, activated NK cells, monocytes, peripheral blood leukocytes, and myeloid cell lines (U937, THP1, K562).As a member of chemokines, CXCL5 has been shown that it is highly expressed in tumors such as liver cancer [10], cholangiocarcinoma [11], colorectal cancer [12], and prostate cancer [13], and regulates tumor proliferation and migration through multiple mechanisms.Zhou Jingxian’s studies have shown that the rs2248374 / rs2549782-AG haplotype of ERAP2 gene may increase the risk of non-small cell lung cancer, while the GT haplotype may be protective, and the ERAP2 gene may be related to [14] in the pathological type of non-small cell lung cancer.

In terms of tumor clinical survival, our study also demonstrated that five key downregulated genes are associated with tumor clinical survival and tumor stage.Among them, downregulation of GART was associated with low survival of KIRC and READ; downregulation of SOD2 was associated with low survival of SKCM, and downregulation of KLRF1 was associated with low survival of HNSC.A positive correlation of GART gene expression levels with tumor stage of LUAD, The GART expression levels were the highest in the LUAD IV stage, It suggests that GART may be involved in LUAD transfer; The SOD2 gene expression levels were correlated with the tumor stage of the SKCM, SOD2 gene expression levels were highest in STAND tumor stage I, The gene expression level was lowest in SOD2 in tumour stage 0; Positive correlation of CXCL5 gene expression levels with the tumor stage of KICH and KIRC, CXCL5 expression was the highest in KICH and KIRC IV stages, It is suggested that CXCL5 may be involved in the transfer of KICH and KIRC.In existing studies, GART has received wide attention as a research hot topic of cancer chemotherapy, and some studies have shown that high GART expression is unfavorable to the prognosis of HCC patients and involves [15] with HCC cell proliferation.Huang showed that high expression in renal transparent cell carcinoma, and its high expression with sex, pathological grade, T stage and N stage, the overall survival of patients with high CXCL5 expression is shorter, indicating that CXCL5 expression is an independent risk factor affecting the prognosis of renal transparent cell carcinoma (P <0.05), CXCL5 gene may be involved in DNA replication, autophagy pathway, PPAR signaling to regulate renal transparent cell carcinoma [16].Liu Shiguang et al study found that the combination of CXCL5 and PD-L1 could stratify the risk degree in colorectal cancer patients and predict the patient prognosis of [17].Yang Gang et al found that miR-27 targeting negatively regulates SOD2 and inhibition of SOD2 expression can reverse inhibit the inhibitory effect of miR-27 on MCF-7 proliferation, migration, and invasion in breast cancer cells, [18].Lu Shizhen et al found that the chemokine CXCL5 promoted the [19] proliferation and migration of cervical cancer cells through the autocrine pathway through the ERK signaling pathway.

Our study also analyzed the mutations and correlation of five key genes in tumors, suggesting that among the five key genes, mutations and amplification are the most common types, and that the mutations are mainly missense mutations.

Based on the above studies, we hypothesized that key downregulated genes for reMI may in some cases lead to tumorigenesis and promote tumor metastasis, and that the expression level of these genes may affect tumor clinical survival and tumor stage.The main limitation of this study is that the number of cases of the chip data set included in this study is small, lack of representativeness and universality, and the later stage should search other databases or conduct multi-center clinical research; second, the lack of further verification of relevant animal or cell experiments in this study.

## 4. Conclusion

In conclusion, we comprehensively elucidated the genomics of reMI after the first MI and its association with tumors.Our findings reveal that key downregulated genes in reMI are associated with tumorigenesis, metastasis, clinical survival, immune infiltration.Key downregulated genes for reinfarction may be used as important biomarkers for the diagnosis and treatment of cancer.

## Data Availability

All data produced in the present study are available upon reasonable request to the authors
All data produced in the present work are contained in the manuscript
All data produced are available online at GEO, TIMER, GEPIA2, cBioPortal, and DAVID

## Acknowledgements

None.

## Authors’ contributions

WZY conceived the study, designed the experiments, collected data, and wrote the manuscript.

## Availability of data and materials

The datasets generated and analyzed during the current study are available from the corresponding author on reasonable request. The raw data could also be obtained from online databases including the GEO, TIMER, GEPIA2, cBioPortal, and DAVID without any restrictions.

